# Comparing rapid micro-induction and standard induction of buprenorphine/naloxone for treatment of opioid use disorder: Protocol for an open-label, parallel-group, superiority, randomized controlled trial

**DOI:** 10.1101/2020.05.22.20106062

**Authors:** James S.H. Wong, Mohammadali Nikoo, Jean N. Westenberg, Janet G. Suen, Jennifer Wong, Reinhard M. Krausz, Christian G. Schütz, Marc Vogel, Jesse A. Sidhu, Jessica Moe, Shane Arishenkoff, Donald Griesdale, Nickie Mathew, Pouya Azar

## Abstract

**Background:** Buprenorphine/naloxone (Suboxone) is a current first-line treatment for opioid use disorder (OUD). The standard induction method of buprenorphine/naloxone requires patients to be abstinent from opioids and therefore experience withdrawal symptoms prior to induction, which can be a barrier in starting treatment. Rapid micro-induction (micro-dosing) involves the administration of small, frequent does of buprenorphine/naloxone and removes the need for a period of withdrawal prior to the start of treatment. This study aims to compare the effectiveness and safety of rapid micro-induction versus standard induction of buprenorphine/naloxone in patients with OUD.

**Methods:** This is a randomized, open-label, two-arm, superiority, controlled trial comparing the safety and effectiveness of rapid micro-induction versus standard induction of buprenorphine/naloxone for the treatment of OUD. A total of 50 participants with OUD will be randomized at one Canadian hospital. The primary outcome is successful induction of buprenorphine/naloxone with low levels of withdrawal. Secondary outcomes are treatment retention, illicit drug use, self-reported drug use behaviour, craving, pain, physical health, safety, and client satisfaction.

**Discussion:** This is the first randomized controlled trial to compare the effectiveness and safety of rapid micro-induction versus standard induction of buprenorphine/naloxone. This study will thereby generate evidence for a novel induction method which eliminates substantial barriers to the use of buprenorphine/naloxone in the midst of the ongoing opioid crisis.

## 1. Background

The opioid crisis is one of the most serious public health issues in North America in recent years. In the United States, over 65,000 people died from using opioids in 2018, and in Canada, nearly 15,000 opioid-related deaths have occurred since 2016 ^1-3^. Opioid-related deaths have surpassed motor vehicle incidents and homicide deaths combined, resulting in decreases in life expectancy in North America ^1,4^.

These deaths are primarily driven by untreated opioid use disorder (OUD), a common disorder affecting millions of individuals worldwide 5. Current North American guidelines strongly endorse opioid agonist treatment (OAT) with buprenorphine/naloxone as the first-line treatment of OUD, because of its superior safety profile and comparable efficacy over other forms of OAT ^6,7^. OAT is associated with reducing mortality, illicit drug use, and improving physical and mental health outcomes ^8^.

Buprenorphine is a partial μ-opioid receptor agonist. The partial agonism results in a ceiling effect on respiratory depression and lower risk for overdose ^9^. To prevent abuse and minimize diversion, buprenorphine is co-formulated with naloxone, an opioid antagonist, in a 4:1 ratio as buprenorphine/naloxone (brand name: SUBOXONE^®^) ^10^. When buprenorphine/naloxone is injected by individuals with OUD, naloxone precipitates withdrawal. When buprenorphine/naloxone is taken as prescribed, that is sublingually, naloxone is poorly absorbed and does not exert any significant clinical effect, leaving the opioid agonist effects of buprenorphine to predominate.

Buprenorphine exhibits a strong biding affinity to the μ-opioid receptor ^9^. When it is introduced in the presence of other opioids with weaker binding affinities, such as heroin, buprenorphine can precipitate withdrawal by displacing other opioids from the receptor. To avoid precipitated withdrawal, the standard method of induction of buprenorphine/naloxone requires patients to be abstinent from other opioids for a set period of time and thus requires patients to be in at least mild withdrawal before its administration ^6,7^. Standard buprenorphine/naloxone induction can thereby be very distressing and time-consuming for patients to tolerate, which can be a barrier for many patients who need this potentially life-saving therapy. Patients who experience significant levels of withdrawal or precipitated withdrawal during the induction process may also be less likely to be retained in treatment ^11^.

To overcome the difficulties of a standard induction method of buprenorphine, a novel induction method, known as micro-induction (also called micro-dosing), is being explored and increasingly employed by many clinicians in Canada, the United States, and other parts of the world ^12-20^. This induction method was first described as the ‘Bernese Method’ in a Swiss case series in 2016 ^13^. The method involves administering buprenorphine at micro-doses once to twice daily, concurrently with the use of a full μ-opioid receptor agonist, to avoid precipitated withdrawal. It did not require the two outpatients to go through withdrawal from opioids prior to induction, and they reached therapeutic doses in ten or more days.

Recently, our team developed a more rapid variation of micro-induction, known as ‘rapid micro-induction,’ which was developed to be primarily used in an inpatient setting. It involves the administration of buprenorphine/naloxone every three to four hours along with the use of a full μ-opioid receptor agonist, resulting in patients reaching therapeutic doses in just three to five days ^16^. The rationale of this ‘rapid’ dosing is based on the hypothesis that buprenorphine reaches peak plasma concentration in approximately an hour ^21^. Rapid micro-induction offers several advantages over a standard induction method - eliminating the abstinence period preceding induction, reducing the risk of precipitated withdrawal, minimizing the symptoms of withdrawal and craving, potentially improving treatment retention, and reducing the time spent in hospital ^16,18^.

Rapid micro-induction and variations of this novel induction method have been extensively described in several case reports and in a recent review, however, they have never been systematically evaluated in a clinical trial ^12-20^. To generate the evidence for this induction method in the midst of the ongoing opioid crisis, we propose the first randomized controlled trial was developed to compare the effectiveness and safety of rapid micro-induction versus standard induction of buprenorphine/naloxone.

## 2. Study design

### 2.1 Overview of study design

This study is a randomized controlled trial comparing the effectiveness and safety of rapid micro-induction versus standard induction of buprenorphine/naloxone. It has received approval from the Research Ethics Board of the University of British Columbia (H19-03254) and is registered in clinicaltrials.gov (NCT04234191). In this open-label superiority trial, eligible patients with OUD will be randomized to either: (a) the rapid micro-induction arm or (b) the standard induction arm (treatment as usual).

The study schema is presented in Figure 1. The study will take place at one site, Vancouver General Hospital (VGH) in Vancouver, British Columbia, Canada. The Complex Pain and Addiction Services (CPAS) is a consulting service in which VGH inpatients with substance use disorders are referred to for treatment and counselling. The study staff at CPAS will pre-screen referrals to determine general eligibility for participation in the study. Patients will then be invited by the study staff to complete the informed consent procedures. Once informed consent is provided, participants will undergo screening procedures to confirm their eligibility.

**Figure 1.**
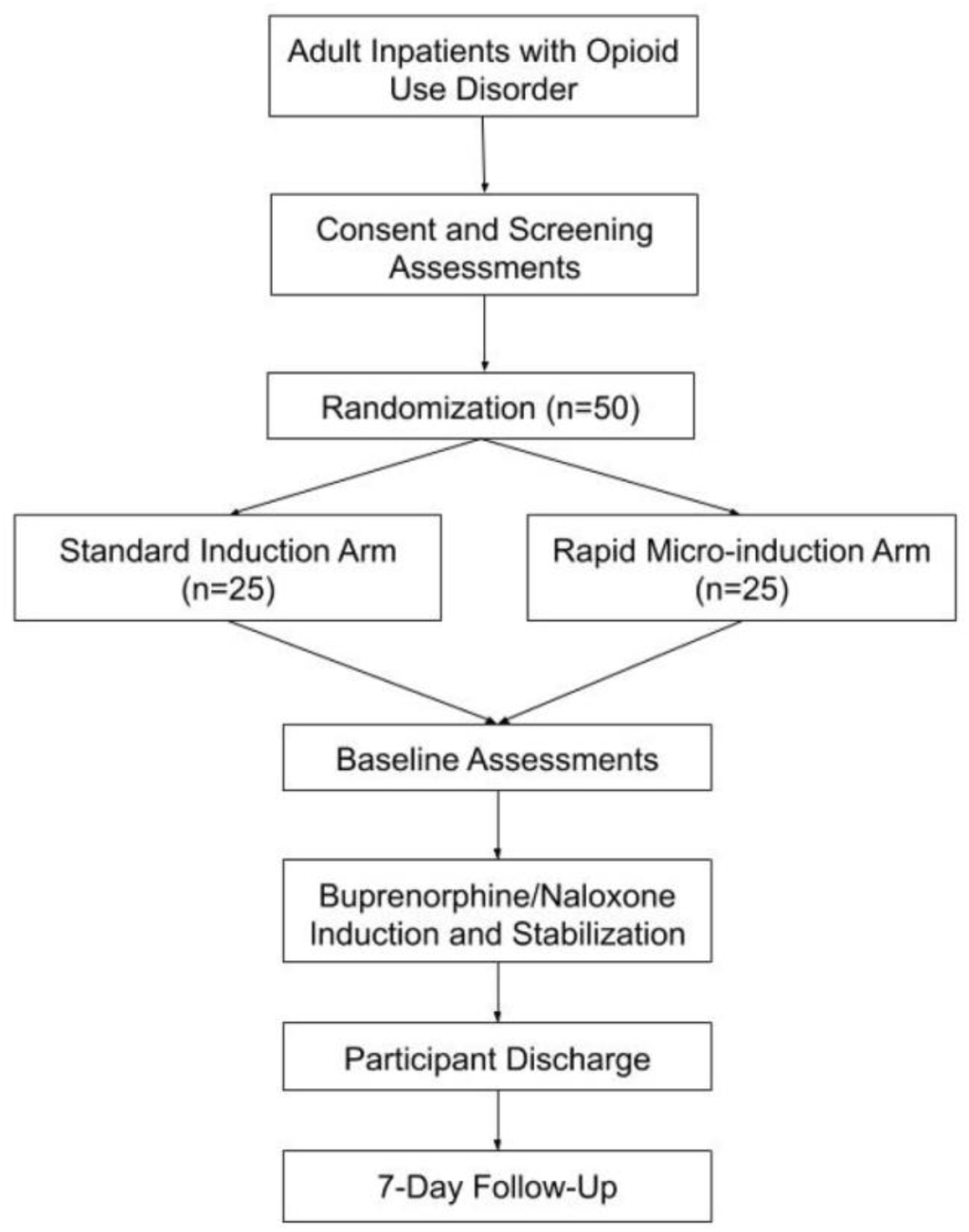
Study schema

Eligible participants will be randomized on an allocation ratio of 1:1 to either of the two arms, using a blocked permuted block design with block sizes of 4 and 6. Randomization will be managed with https://www.sealedenvelope.com. Once randomized, participants will complete baseline assessments and be followed for 7 days. Towards the end of the intervention period, the physician in charge will inform all participants about follow-up treatments that are available to them. Participants will receive follow-up with their community addictions physicians and/or the CPAS outpatient clinic.

### 2.2 Study objectives

The primary objective is to compare rapid micro-induction versus standard induction on the successful induction of buprenorphine/naloxone with low levels of withdrawal in patients with OUD. The secondary objectives are to evaluate treatment retention, illicit drug use, self-reported drug use behaviour, craving, pain, physical health, safety, and client satisfaction.

### 2.3 Study population

Inclusion Criteria

- Opioid use disorder as confirmed by DSM 5 diagnostic criteria
- Individuals seeking opioid agonist treatment (OAT)
- 19 years of age or older
- Willingness to comply with study procedures
- Provide written informed consent to participate in the study
- If female and of childbearing potential, agree to use an effective method of birth control approved by the study investigators throughout the study

Exclusion Criteria

- Diagnosis of severe medical or psychiatric conditions contraindicated for buprenorphine/naloxone and/or hydromorphone treatment
- Anticipated deterioration of health due to discontinuation of medications that are contraindicated with buprenorphine/naloxone and/or hydromorphone
- Positive pregnancy test for females of childbearing potential
- Not experiencing mild to moderate opioid withdrawal after the last dose of methadone
- Positive urine test for methadone
- Known allergy or sensitivity to buprenorphine/naloxone and/or hydromorphone
- Anticipation that the patient may need to initiate pharmacological treatment during the trial that is deemed unsafe by the study physician or could prevent study completion
- Unwilling or unable to use an effective method of birth control approved by the study investigators throughout the study

## 3. Study treatments

The rapid micro-induction arm will involve the administration of buprenorphine/naloxone and hydromorphone, while the standard induction arm will involve the administration of only buprenorphine/naloxone.

### 3.1 Buprenorphine/naloxone

Buprenorphine/naloxone (brand name: SUBOXONE^®^) is the recommended first-line option for the treatment of OUD in Canada and France, and an increasingly popular choice in a number of countries such as the United States and England ^7,22-24^. It will be administered in the form of a sublingual tablet.

### 3.2 Hydromorphone

Hydromorphone is an opioid medication used for managing pain, craving, and withdrawal in this study. The opioids the patients are using will be rotated to hydromorphone. Hydromorphone will be administered as needed to meet the patient’s opioid requirements. It will be administered orally via tablet form; or administered intravenously, subcutaneously, or intramuscularly via liquid form. The route of administration will be determined by the physician in charge in consultation with the patient.

### 3.3 Rapid micro-induction arm

The titration schedule for the rapid micro-induction arm is described in Table 2. This titration schedule has also been termed as a ‘72-hour rapid micro-induction’ because the induction is completed by the end of Day 3. Induction is considered to be completed when patients have received a total daily dose of ≥ 12mg of buprenorphine/naloxone, as 12mg is considered the minimum effective dose according to the product monograph of SUBOXONE^®^ ^25^. On the fourth (last) day of treatment, the dose is consolidated to once daily dosing.

**Table 2.** Rapid micro-induction titration schedule

|  | Buprenorphine/Naloxone* |  |  | Hydromorphone** |  |
| --- | --- | --- | --- | --- | --- |
|  | Dosing | Total Daily Dose |  | Dosing | Total Daily Dose |
| Day 1<br>(0h-24h) | 0.5 mg SL<br>Q3H | 4mg |  | 1 to 16 mg<br>PO/SC/IV/IM<br>Q1-3H PRN and<br>titrate to effect<br>(start at the lower<br>end of dosing<br>range)<br>For 1<sup>st</sup> dose –<br>Max 8mg PO, or<br>4mg SC/IV/IM | Max 96 mg<br>PO/SC/IV/IM |
| Day 2<br>(24h-48h) | 1 mg SL Q3H | 8mg |  | 1 to 16 mg<br>PO/SC/IV/IM<br>Q1-3H PRN and<br>titrate to effect<br>(start at the lower<br>end of dosing<br>range) | Max 96 mg<br>PO/SC/IV/IM |
| Day 3<br>(48h-72h) | 12 mg SL<br>once daily<br>AND<br>1 to 4 mg SL<br>Q3H PRN<br>Max 12 mg | 12mg;<br>Max 24mg<br>including PRN |  | Discontinued |  |
| Day 4<br>(72h-96h) | Consolidate<br>Day 3 total<br>dose to once<br>daily dosing | Max 24mg |  | Discontinued |  |
| *Expressed as milligrams of buprenorphine in a buprenorphine/naloxone SL tablet<br>**The opioids the patients are using will be rotated to hydromorphone. Hydromorphone will be administered as needed to meet the patient’s opioid requirements. Hold if sedated, RR <12, or O2 saturation <92%.<br>SL, sublingual. Q_H, every _ hour. PRN, as needed. PO, by mouth. SC, subcutaneous. IV, intravenous. IM, intramuscular. |  |  |  |  |  |

### 3.4 Standard induction arm

The standard induction titration schedule is described in Table 3. It has been written in accordance to the ASAM Practice Guidelines and the product monograph of SUBOXONE^®^ sublingual tablet ^6,25^. Day 1 is initiated when participants score 11 or above on the Clinical Opiate Withdrawal Scale (COWS), and when they have been abstinent from short-acting opioids for at least 6-12 hours or from long-acting opioids for 24-72 hours. Induction is considered to be completed when patients have received a total daily dose of ≥ 12mg of buprenorphine/naloxone. On the third (last) day of treatment, the dose is consolidated to once daily dosing.

**Table 3.** Standard induction titration schedule

|  | Buprenorphine/Naloxone* |
| --- | --- |
|  | Dosing |
| **Day 1\*\***<br>**(0h-24h)** | Start with 2 to 4 mg SL.<br>If 60-90 minutes have passed without the onset of withdrawal symptoms: additional dosing can be done in increments of 2 to 8 mg SL.<br>Suggested total dose target for Day 1 is 8 to 12 mg SL |
| **Day 2**<br>**(24h-48h)** | Start with dose equal to the total amount of buprenorphine/naloxone administered on Day 1.<br>Titrate in increments or decrements of 2 to 8mg to a level that holds the patient in treatment and suppresses opioid withdrawal effects is guided by reassessment of the clinical and psychological status of the patient.<br>Suggested total daily dose for Day 2 is 12 to 16mg SL.<br>Max total daily dose is 24mg SL. |
| **Day 3**<br>**(48h-72h)** | Consolidate Day 2 total dose to once daily dosing.<br>Suggested total daily dose for Day 3 is 12 to 16mg SL.<br>Max total daily dose is 24mg SL. |
| *Expressed as milligrams of buprenorphine in a buprenorphine/naloxone SL tablet<br>**Day 1 is initiated when patient is COWS Score $\geq 11$; and at least 6-12 hours after last use of short-acting opioids or 24-72 hours after last use of long-acting opioids<br>SL, sublingual. COWS, Clinical Opiate Withdrawal Scale. |  |

### 3.5 Medical management

In both the standard induction and rapid micro-induction arms, residual withdrawal symptoms will be managed as per ASAM guidelines: clonidine may be used at doses of 0.1 - 0.3 mg every 6 – 8 hours, with a maximum dose of 1.2 mg daily (American Society of Addiction Medicine, 2020). Other non-narcotic medications targeting specific opioid withdrawal symptoms can also be used as per ASAM guidelines: benzodiazepines for anxiety, loperamide or bismuthsalycilate for diarrhea, acetaminophen or non-steroidal anti-inflammatory medications (NSAIDs) for pain, zopiclone for insomnia, and ondansetron for nausea.

All participants will receive routine motivational interviewing, behavioural-based psychoeducation, and supportive psychotherapy provided by the CPAS team based on the individual’s need. The type, duration, and reason for all the psychological interventions and medications provided will be documented in the case report form (CRF).

## 4. Outcomes and assessments

The timeline of assessments is shown in Table 4. It should be noted that there is an additional day of assessments conducted with the experimental arm, because the experimental intervention (rapid micro-induction) takes one day longer than the treatment-as-usual intervention (standard induction).

**Table 4.**
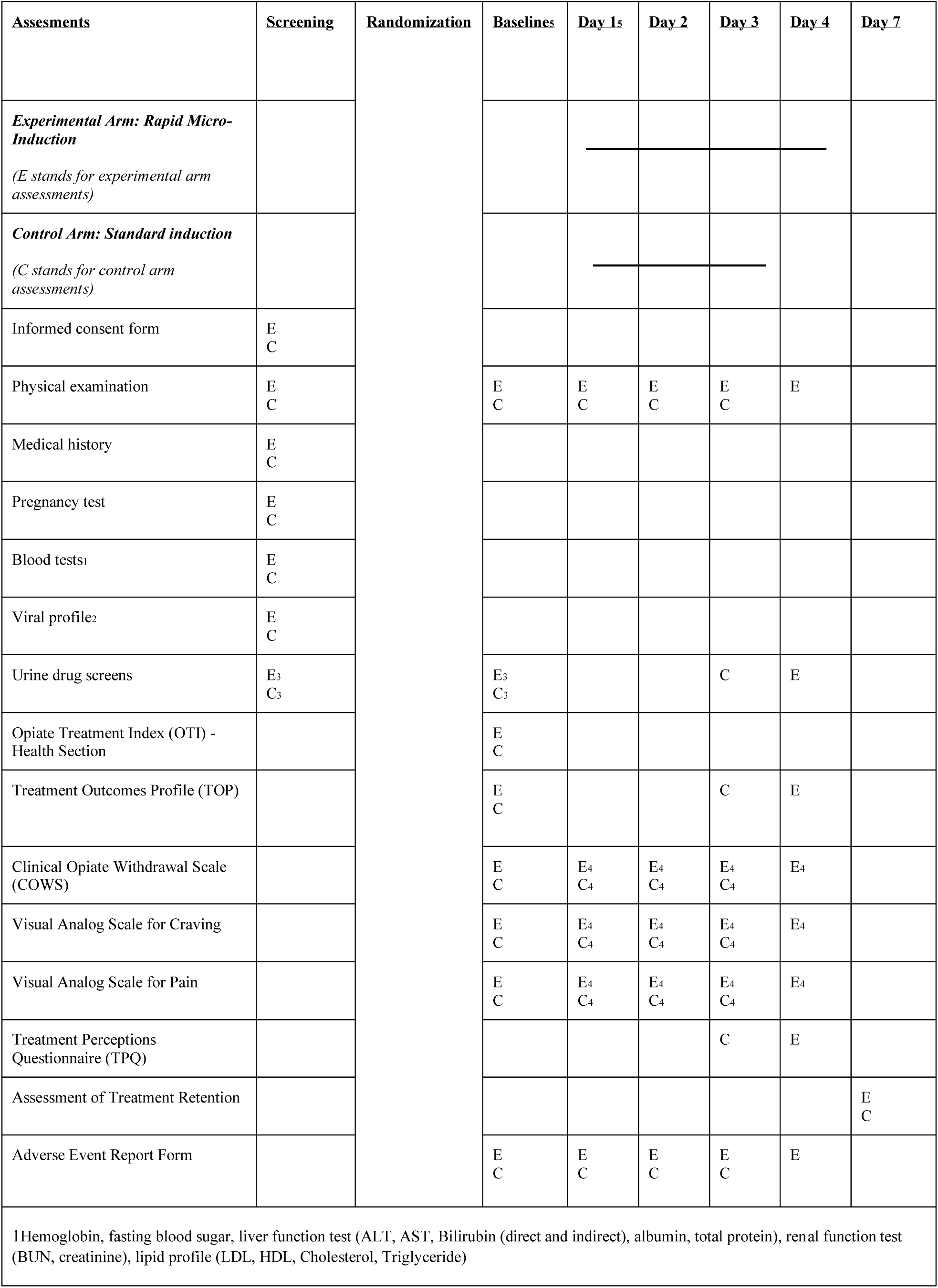
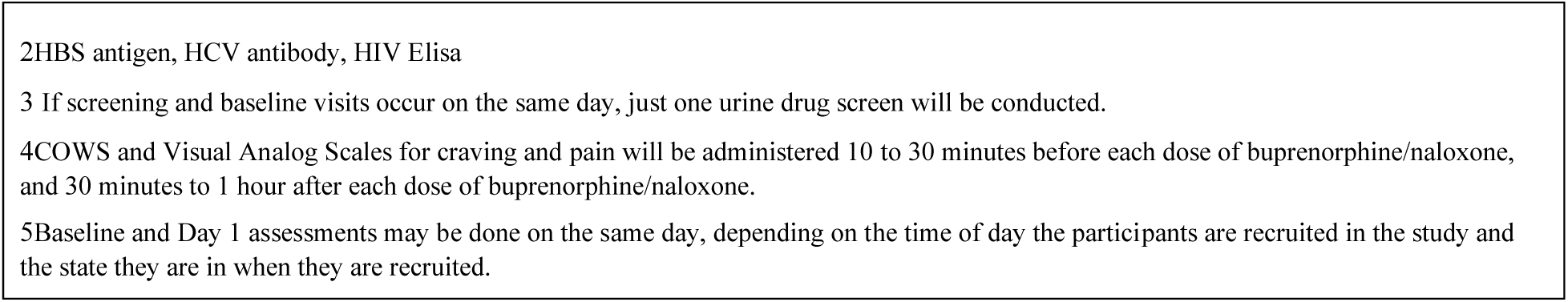
Timeline of assessments

### 4.1 Primary outcome

The primary outcome is successful induction of buprenorphine/naloxone with low levels of withdrawal. This is defined as the following: participants who remain in treatment until they have received a total daily dose of ≥ 12mg of buprenorphine/naloxone (successful induction), and score ≤ 12 on the COWS (low levels of withdrawal) from the time of randomization to when they reach that dose ^26^. The COWS will be administered at baseline, days 1 to 4 of the experimental arm, and days 1 to 3 of the control arm – specifically, 10 to 30 minutes before each dose of buprenorphine/naloxone, and 30 minutes to 1 hour after each dose of buprenorphine/naloxone

### 4.2 Secondary outcomes

The secondary outcomes are treatment retention, illicit drug use, self-reported drug use behaviour, craving, pain, physical health, safety, and client satisfaction.

Treatment retention will be assessed by buprenorphine/naloxone prescription pick-up on day 7.

Illicit drug use will be assessed by urine drug screens, which will analyze for the presence of cocaine, opioids including methadone, buprenorphine, hydromorphone, oxycodone, morphine, heroin, and fentanyl, benzodiazepines, amphetamines, and methamphetamine. Urine will be collected at screening, baseline, day 4 of the experimental arm, and day 3 of the control arm.

Self-reported drug use behaviour will be assessed by the Treatment Outcome Profile (TOP) at baseline, day 4 of the experimental arm, and day 3 of the control arm. The TOP is a 20-item instrument designed to assess and monitor substance misuse by measuring four different domains (substance use, health, crime and social functioning) and includes thirty-eight frequency, rating scale and period prevalence measure ^27^.

Craving and pain will be measured by visual analog scales (VAS) at baseline, days 1 to 4 of the experimental arm, and days 1 to 3 of the control arm. The VAS for craving and pain will be administered 10 to 30 minutes before each dose of buprenorphine/naloxone, and 30 minutes to 1 hour after each dose of buprenorphine/naloxone. The VAS presents the participant a rating scale which represents the spectrum of pain and craving: the left end indicates no pain or craving while the right end indicates extreme pain and craving. Participants mark the 10 centimeter rating scale at a location on along the spectrum which most accurately reflects the craving or pain they are experiencing ^28^.

Physical health will be assessed by the health section of the Opiate Treatment Index (OTI) at baseline. The OTI is a structured interview designed to provide a measure of the effectiveness of drug treatments, by measuring six outcomes: drug use, HIV risk-taking behaviour, social functioning, criminality, health status, and psychological functioning ^29^. Only the health section of the OTI will be used; the health section is composed of items addressing signs and symptoms in major organ systems and injection-related health problems.

Safety will be assessed by the appearance of adverse events (AEs) and serious adverse events (SAEs), which will be recorded on the CRF. AEs and SAEs are defined in Section 6.1 Safety.

Client satisfaction will be assessed by the Treatment Perceptions Questionnaire (TPQ) on day 4 of the experimental arm and day 3 of the control arm. The TPQ is a 10-item scale which assesses the satisfaction of patients in addiction treatment program, examining two areas: perception of clients towards the nature and extent of their contact with the program staff (5 items), and aspects of the treatment service and its operation and rules and regulation (5 items) ^30^.

## 5. Sample size and power calculation

The sample size calculation for the binary primary outcome is based on testing for superiority in a parallel group clinical trial. As only case reports have been published on rapid micro-induction, we expect a success rate of 95% in the experimental arm based on the opinion of two addiction psychiatry experts familiar with the method. We expect a success rate of 10% in the control arm, as most participants in the arm are anticipated to experience moderate to higher levels of withdrawal, which is defined as having a COWS score of ≥ 13. A difference of such a magnitude, 85%, is deemed clinically meaningful. Using G*Power 3.1 software, the minimum required sample size to power a Fisher’s exact test to detect this difference between the two arms with a type I error of 0.05 and a type II error of 0.1 will be 12 (6 in each arm). Adjusting for an attrition rate of 10% (participants with incomplete COWS data, participants who have discontinued the treatment they were randomized, and participants who have discontinued both treatment and data collection procedures), the required sample size is 14. We aim for a sample size of 50 (25 each arm), as a larger sample size is not feasible due to cost and constraints.

## 6. Safety, treatment discontinuation, and study discontinuation

### 6.1 Safety

Safety will be assessed by the appearance of adverse events (AEs) and serious adverse events (SAEs). AEs and SAEs will be monitored throughout the study. An AE is defined as any untoward medical occurrence in a participant, administered a study intervention, which does not necessarily have a causal relationship with this intervention. A SAE is defined as any untoward medical occurrence meeting one of the following criteria at any dose: results in death, is life-threatening, requires inpatient hospitalization or prolongation of existing hospitalization, results in persistent or significant disability or incapacity, or is a congenital anomaly or birth defect. All AEs and SAEs will be documented accordingly on the adverse event report form, and will be reported to the clinical trial’s Data Safety and Monitoring Board (DSMB). All SAEs will be reported to the Sponsor-Investigator and Health Canada.

### 6.2 Treatment discontinuation and study discontinuation

Participants are free to discontinue the treatment arm they were randomized to (treatment discontinuation), or the treatment they were randomized to and any data collection procedures (study discontinuation). Discontinuation may occur at any time without participants having to provide any reason and without prejudice to their medical care. Discontinuation from treatment or study may occur in the following circumstances, but not limited to: participant’s request, severe adverse reactions/events and/or other safety reasons, violence against treatment team without convincing evidence of mental illness like psychosis or delirium, and criminal behaviour with resulting imprisonment during the study period. All discontinuations from treatment or study will be documented on the CRF. Such participants will be considered failures of the primary outcome.

If participants discontinue the treatment they were randomized to (treatment discontinuation), they will be offered the other parallel treatment. This will be determined by the physician in charge in consultation with the patient. In order to reduce the amount of missing or incomplete data from such participants, research staff will continue to collect data from the participants if they allow them to do so.

If participants discontinue from the study (study discontinuation), the research staff will not collect data from them, and CPAS physicians will follow-up with appropriate treatment options.

## 7. Data analysis

All analyses will be conducted with both intention-to-treat (ITT) and per-protocol (PP) methods. Participants who discontinue the treatment arm they were randomized to (includes switches to the other arm) will be considered as failures of the primary outcome.

### 7.1 Primary outcome

The primary outcome will be assessed in a binary fashion: participants need to fulfill both criteria (successful induction and low levels of withdrawal) in order to be successful with the primary outcome. Outlier COWS scores will be checked and confirmed with the clinical records on an ongoing basis by the study coordinator. Effort will be made by the study team to avoid missing COWS scores during the study. In case of missing scores, multiple imputation will be used to impute the missing variables, and sensitivity analysis will be performed by once assigning failure to all the missing COWS score and then success to them, then describing and explaining the impact on the results. Fisher’s exact test will be used to compare two groups with a significance level set at 0.05. To demonstrate the effect size, we will use both unadjusted and adjusted odds ratios with 95% confidence intervals using logistic regression. In the latter, effect size will be adjusted for age, gender, and baseline COWS score in addition to allocated arm. Adjustment for baseline covariates will improve the sensitivity of the comparison.

### 7.2 Secondary outcomes

Secondary outcomes will be compared between two arms using Fisher’s exact, Wilcoxon-Mann Whitney, and interaction terms from Linear Mixed Models for binary, interval, and repeated measures, respectively.

## 8. Current status of the study

As of February 2020, the study has received approval from the Ethics Board of the University of British Columbia and Health Canada to use buprenorphine/naloxone off-label in the trial. The study site, the Complex Pain and Addiction Services (CPAS) at Vancouver General Hospital (VGH) in Vancouver, British Columbia, Canada, has been preparing for recruitment. As per discussion with experts working with CPAS, it is estimated that the recruitment rate will be 12 participants per month.

## 9. Discussion

This is the first study to compare the safety and effectiveness of rapid micro-induction and standard induction of buprenorphine/naloxone for the treatment of OUD. This study was initiated in response to the lack of research evaluating novel buprenorphine/naloxone induction protocols. While buprenorphine/naloxone has been widely accepted as a treatment method for OUD due to its superior safety profile compared to other OAT options, there are still several barriers that have prevented its widespread use. One major barrier is that the standard induction of buprenorphine/naloxone requires patients to be in a period of opioid withdrawal prior to starting treatment. Patients may be fearful of experiencing withdrawal associated with the standard induction protocol, which in turn may affect their retention in treatment ^11^. This may also lead patients to try other forms of OAT with less favourable safety profiles, such as methadone and slow-release morphine ^7^. Thereby, ensuring a safer and more comfortable induction process for patients may improve treatment retention and decrease their risk of overdose.

The use of alternative induction protocols, such as rapid micro-induction, have consequently been utilized to address the concerns with the standard induction process. Buprenorphine/naloxone rapid micro-induction confers many benefits over a standard induction method, as it can minimize withdrawal and craving symptoms, reduce the risk of precipitated withdrawal, and length of induction ^16,18^. Anecdotally and according to recent case reports and a review, clinicians have had much success with rapid-micro-induction, and the method has entered routine practice at some hospitals and clinics across Canada, the United States, and Europe ^12-20^.

The open-label design of this clinical trial may introduce the risk of potential bias. However, it is not feasible to blind participants or researchers due to the nature of the interventions. The study may also benefit from a large sample size and a longer duration of follow-up, but such changes are not possible due to resource and cost constraints. Despite these limitations, the evaluation of the primary and secondary outcomes will greatly contribute to our understanding of rapid micro-induction. As the opioid crisis in North America continues, the results derived from this clinical trial will generate the first major body of clinical evidence on the effectiveness and safety of rapid micro-induction, a novel and patient-centered induction approach which could immensely improve the accessibility of buprenorphine/naloxone for patients with OUD.

## Data Availability

Not applicable.

## Acknowledgements

We thank Ehsan Moazen-Zadeh (Icahn School of Medicine at Mount Sinai), Fiona Choi (University of British Columbia), Mostafa Mamdouh (University of British Columbia), Kimia Ziafat (University of British Columbia), and Kiana Kianpoor (University of British Columbia) for helpful discussions.

